# Sero-prevalence of anti-SARS-CoV-2 antibodies in Chattogram Metropolitan Area, Bangladesh

**DOI:** 10.1101/2022.02.09.22270717

**Authors:** Jahan Ara, Md. Sirazul Islam, Md. Tarek Ul Quader, Anan Das, F. M. Yasir Hasib, Mohammad Saiful Islam, Tazrina Rahman, Seemanta Das, M.A. Hassan Chowdhury, Goutam Buddha Das, Sharmin Chowdhury

## Abstract

**Background:** Seroprevalence studies of coronavirus disease 2019 (COVID-19) assess the degree of undetected transmission in the community. Different groups, such as healthcare workers (HCWs), garment workers, and others, are deemed vulnerable due to their workplace hazards and immense responsibility.

**Purpose:** The present study was conducted to estimate the seroprevalence of anti-SARS-CoV-2 antibody (IgG) and its association with different explanatory variables. Further, the antibody was quantified to assess the increasing or decreasing trend over different intervention periods and according to other factors.

**Methodology:** This cross-sectional study observed health workers - doctor, nurse, hospital staff, etc. in and outpatients (non-COVID-19) and garments workers of Chattogram metropolitan area (CMA, N=748) from randomly selected six government and private hospitals and two garment factories. Study subjects were included upon written consent, fulfilling specific inclusion criteria. Venous blood was collected following standard aseptic methods. Qualitative and quantitative ELISA was used to identify and quantify antibodies (IgG) in serum samples. Descriptive, univariable, and multivariable statistical analysis was performed.

**Results:** Overall seroprevalence was estimated as 66.99% (95% CI: 63.40%-70.40%). Seroprevalence among HCWs, in and outpatients, and garments workers were 68.99 % (95% CI: 63.8%-73.7%), 81.37 % (95% CI: 74.7%-86.7%), and 50.56 % (95% CI: 43.5%-57.5%), respectively. Seroprevalence was 44.47 % (95% CI: 38.6%-50.4%) in the non-vaccinated population while it was significantly (*p* <0.001) higher in the population receiving the first dose (61.66 %, 95% CI: 54.8%-68.0%) and both (first and second) doses of vaccine (100%, 95% CI: 98.4%-100%). The mean titer of the antibody was estimated as 255.46 DU/ml and 159.08 DU/ml in the population with both doses and one dose of vaccine, respectively, compared to 53.71 DU/ml of the unvaccinated population. A decreasing trend in the titer of antibodies with increasing time after vaccination was observed.

**Conclusions:** Seroprevalence and mean antibody titer varied according to different factors in this study. The second dose of vaccine significantly increased the seroprevalence and titer, which decreased to a certain level over time. Although antibody was produced following natural infection, the mean titer was relatively low compared to antibody after vaccination. This study emphasizes the role of the vaccine in antibody production. Based on the findings, interventions like continuing extensive mass vaccination of the leftover unvaccinated population and bringing the mass population with a second dose under a third dose campaign might be planned.

## Introduction

Chattogram, the port city of Bangladesh, is classified as a high-risk zone for SARS-CoV-2 contact transmission and is one of the most crowded economic and trading centers [**1**]. On April 3, 2020, Chattogram city witnessed its first Coronavirus Disease 2019 (COVID-19) positive case [**2**], followed by the first death on 9 April [**3**]. The disease can manifest itself in various ways, from asymptomatic and minor upper respiratory symptoms to severe pneumonia and acute respiratory distress syndrome [**4**]. While nucleic acid amplification, such as polymerase chain reaction (PCR), is the gold standard for diagnosing acute SARS-CoV-2 infection and is widely recommended, the antibody-based approach improves diagnosis accuracy by capturing asymptomatic testing and recovered infections [**5**].

During an infectious disease outbreak, seroprevalence investigations are crucial in revealing undetected infection in the population and preventing post-pandemic reappearance [**6**]. Determining the actual burden of infection is also vital for epidemic forecasting and response planning. Seroprevalence studies are potent in identifying the number of undiagnosed missing cases with mild or no symptoms or who cannot undergo testing that may contribute significantly to the transmission [**7–11**]. Further, seroprevalence studies estimate the susceptible population in a community. A current investigation discovered that up to 23% of the patients diagnosed with COVID-19 from December 2020 to February 2021 in Bangladesh were asymptomatic [**12**]. Thus, antibody testing could be crucial to determine the actual SARS-CoV-2 exposure rates since PCR only identifies the viral nucleic acid in individuals with existing symptoms [**13**].

According to numerous research, seropositivity fluctuates considerably depending on parameters such as location and time **[7, 14**]. Antibody titers reach their peak one month after the onset of symptoms, and their levels are directly proportional to the severity of the illness [**15**]. Titers continue to fall after that, with IgM and IgA titers falling fast and IgG titers falling more slowly [**16**]. However, a greater understanding of antibody responses to SARS-CoV-2 after natural infection might aid in the development of more successful vaccination strategies in the future. Bangladesh started administering COVID-19 vaccinations on January 27, 2021, and mass immunization commenced on February 7, 2021 [**17, 18**]. As of December 21, 2021, 50.27% of the target population had received the first dose, and 34.60% received the second dose [**19**]. Bangladesh has already started administering third doses to senior persons aged 60 and up, people with comorbidities, and frontline workers. [**20**]. According to a web-based anonymous cross-sectional survey conducted among the general Bangladeshi population between January 30 and February 6, 2002, 61.16% of respondents were inclined to accept/take the COVID-19 vaccine [**21**]. However, vaccination coverage and seroprevalence among the general public must be investigated nationwide to know the herd immunity.

In the COVID-19 pandemic, HCWs are facing immense challenges worldwide. Occupational exposures among HCWs have been documented in numerous nations as worrying [**22**]. Likewise, COVID-19 has had a significant impact on the healthcare system of Bangladesh. According to the latest data from the Bangladesh Medical Association, between March 8, 2020, and November 11, 2021, 9455 HCWs, including physicians, nurses, and other staff, were infected with COVID-19, as well as 188 doctors died as a result [**23**]. Front liners directly involved in diagnosing, treating, and caring for COVID-19 patients are at risk of physical and psychological distress [**24–29**]. Similarly, workers in the garment industry confront different problems in the workplace all around the world. According to the Bangladesh Garment Manufacturers and Exporters Association (BGMEA), 4500 garment companies employ over 4.5 million people or nearly 2.5 percent of the country’s entire population [**30**]. The bulk of the industries operate with limited space, making it challenging to enforce physical distancing norms [**31**]. SARS-CoV-2 transmission might be exacerbated by crowded workplaces, transportation, and lack of physical distancing [**32**]. Hence, it is necessary to put in place measures including risk management in the workplace, vulnerable employee care, the development of an occupational surveillance system, and vaccination policy administration to address the COVID-19 issues [**33, 34**]. Thus, knowing the true seroprevalence both in the risk groups and community might assist in planning interventions efficiently.

In this study, we reported population-based SARS-CoV-2 seropositivity among HCWs, indoor and outdoor patients of various government and private hospitals, and garment workers of CMA, as determined by enzyme-linked immunosorbent assay (ELISA). Moreover, we measured the antibody titer, and both outcomes (seropositivity and antibody titer) were tested to know the association of different factors.

## Materials and Methods

### Study design and setting

From February to September 2021, we conducted a cross-sectional population-based study among HCWs (e.g., doctors, nurses, hospital staff, ward boy, and cleaner), garment workers, and indoor and outdoor patients (non-COVID-19) of six government and private hospitals each, and two garment factories in CMA. All hospitals belonging to the study area were stratified according to their affiliation status; government and private. From each stratum, six hospitals were randomly selected. Sample size was calculated considering the following parameter: 0.65 proportion, 5% margin of error, 95% confidence limit and design effect 2. Each organization’s human resources department provided a list of personnel. Following a simple random sampling technique, samples were collected from a total of 748 respondents.

We interviewed participants to collect information after receiving written consent. Answering a questionnaire and taking blood to test SARS CoV-2 antibodies were part of the study procedure. Our study followed a World Health Organization protocol for population-level COVID-19 antibody testing [**35**]. The questionnaire included sociodemographic details and factors hypothesized to be associated with seropositivity. Participants were included in the study based on several inclusion criteria.

Inclusion criteria

- **Asymptomatic**: Only an asymptomatic group was included to ensure the presence of antibodies. Participants had no COVID-19 related clinical signs, e.g., fever, coughing, runny nose, sore throat, dyspnea, shortness of breath, aches and pain at the time of sample collection
- **In case of having past confirmed COVID-19 status (by Rt PCR**):
  i. Participants who had already passed at least 28 days after a negative Rt-PCR test.
  ii. Participants who did not take a repeated test to ensure negativity had passed at least 42 days after the first COVID-19 test.

Besides, persons under 18 were excluded, as were those with an incomplete questionnaire.

### Baseline blood collection and processing

Heparinized blood specimens (6mL) were collected and transported to the clinical pathology laboratory (CPL) of Chattogram Veterinary and Animal Sciences University (CVASU) within three hours of collection. The serum was separated to evaluate the IgG antibody and kept at −20 °C until serological investigation.

### Serological test examination

Antibody was determined by a commercial qualitative assay using COVID-19 IgG ELISA test (Beijing Kewei Clinical Diagnostic Reagent Inc., China; Ref: 601340) as per the manufacturer’s instructions. The assay is an enzyme-linked immunoassay (ELISA) that detects IgG against the SARS-CoV-2. An index (Absorbance/Cutt-off) of <1 was interpreted as negative, 0.9 to 1.1 as borderline (retesting of these specimens in duplicates was done to confirm results), and ≥1 index as positive. Per the manufacturer, the sensitivity and specificity of the assay for IgG are 93.8% and 97.3%, respectively. Positive and negative controls were included in all assay batches. Repeated testing using the same specimen yielded the same interpretation.

The concentration of IgG antibodies was determined by SARS-CoV-2 S1-RBD IgG (DiaSino® Laboratories Co., Ltd. Zhengzhou, China, Ref: DS207704), which is based on enzyme-linked immunoassay for the quantitative detection of IgG antibodies. The assay’s sensitivity and specificity for IgG quantification, according to the manufacturer, are 98.41% and 98.02%, respectively. Quantitative results were calculated as a ratio of the extinction of the control or tested specimen over the extinction of the calibrator. Results were reported in standardized units for the quantitative kits that included six calibrators to quantify the antibody concentration (i.e., DiaSino units/mL). A value of <10 DU/mL was considered negative, and values >10 DU/mL were positive.

### Data management

The linearity of the quantitative variables was evaluated by categorizing them into four categories using quartiles as cut-off values. Logistic regression analysis was conducted on the categorized variables, and parameter estimates were observed for an increasing or decreasing trend. In case of linear increase or decrease in the parameter estimates, linearity in the quantitative variable was assumed and used without modification. In the case of nonlinearity, a quartile was used to categorize it. However, some quantitative variables were categorized considering research interest. For instance, the number of days between the first dose of vaccine and quantification of antibody titer was categorized as ‘after one month’ and ‘after two months’ and between the second dose of vaccine and quantification of antibody titer was categorized as ‘after two months’, ‘after four months’ and ‘after six months’. The number of days between the vaccination and the antibody titer was achieved from the date of vaccination and sample collection. The prevalence estimates were adjusted with the test kit performance (sensitivity and specificity), and the adjusted prevalence was denoted as true prevalence.

### Data analysis

In the study period, a total of 748 qualitative and quantitative test results were included in the analysis. To evaluate the correlation and collinearity in the categorical and quantitative variables, Cramer’s V test, Spearman correlation coefficient, Chi-square test, t-test or ANOVA, where appropriate, was used. Variables with a significant association or a Spearman correlation coefficient above 0.4 were regarded as correlated. The effects of different potential explanatory variables on the binary outcome - presence/absence of anti-SARS-CoV-2 antibody, was evaluated using univariable and followed by multivariable logistic regression models. To select the final multivariable model, all variables with a significant p-value in the univariable models were included in a model and a manually conducted backward selection strategy was followed by deleting one variable at a time with the highest P-value. Interactions between all explanatory variables (2 ways) were evaluated in the final model. Effect of variables on the mean titer of the antibody was assessed by t-test and one way ANOVA. P-values <0.05 were considered as significant throughout the analysis. STATA-IC 13 (StataCorp, California, USA) and GraphPad Prism 7.00 for Windows (GraphPad Software, La Jolla, California, USA) were used for statistical analyses and visualization.

### Ethical approval and informed consent

Institutional ethical approval was taken from the authorized committee of Chattogram Veterinary and Animal Sciences University (CVASU), Bangladesh [CVASU/Dir(R&E) EC/2020/212(1)].

## Results

### Sero-prevalence of SARS-CoV-2 infection

SARS-CoV-2 IgG antibodies were detected in 498 (66.99%) of 748 individuals **(Table 1)**. Prevalence of anti-SARS-CoV-2 antibody (IgG) in different donor types along with vaccination percentage is shown in **Figure 1**.

**Table 1:** Prevalence estimation in CMA

| Table 1: Prevalence estimation in CMA |  |  |  |  |
| --- | --- | --- | --- | --- |
| Anti-SARS CoV-2 antibody | Total population | Unadjusted seroprevalence, % (95% CI) | Test performance adjusted seroprevalence % (95% CI) | Known positives (RT-qPCR positive) (%) |
| Present | 498 | 66.58 (63.1-70.0) | 66.99 (63.40-70.40) | 91 (80.53) |
| Absent | 250 | 33.42 (30.1-36.9) | 32.60 (29.20-36.19) | 22 (19.47) |

**Figure 1:**
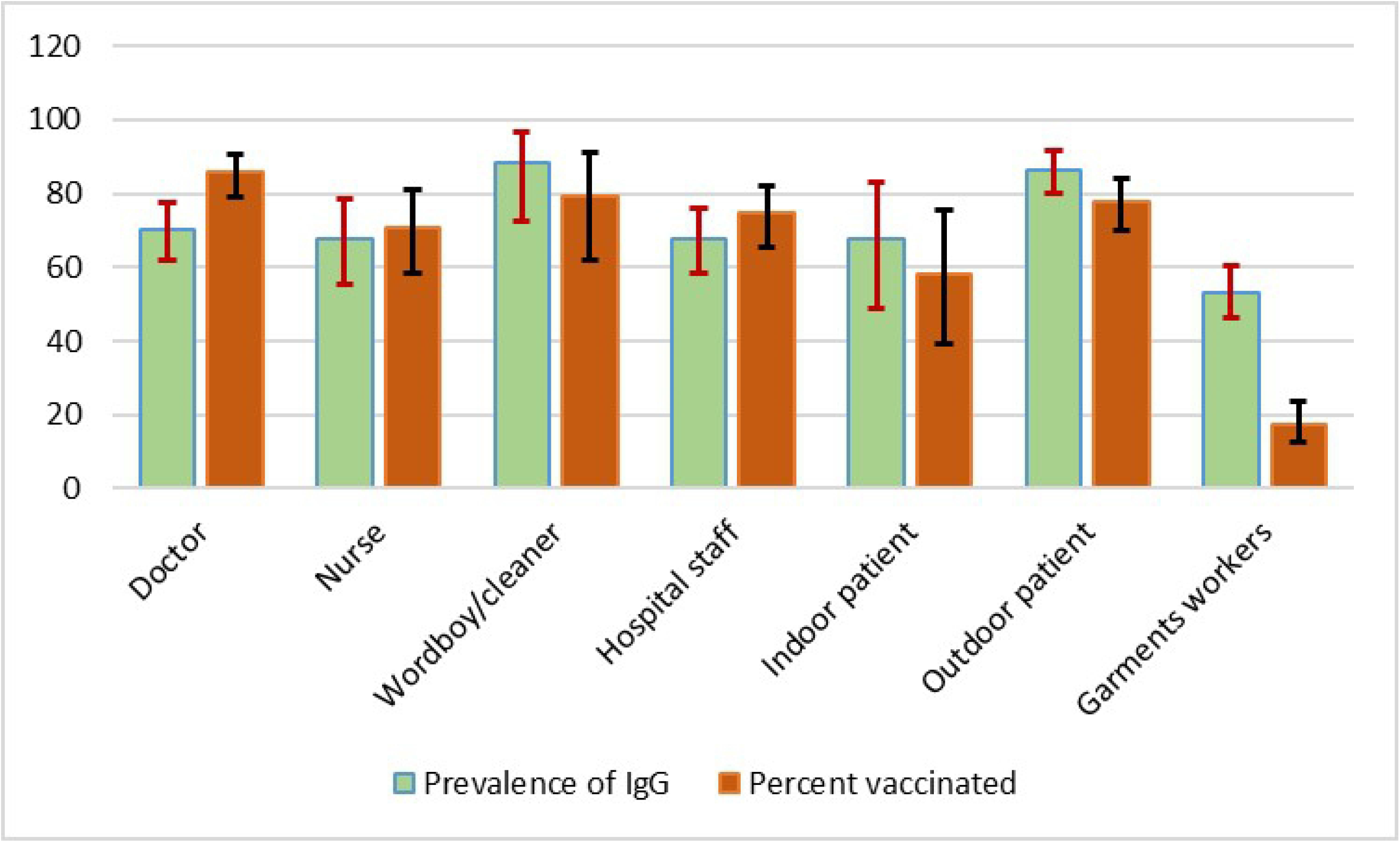
Prevalence of anti-SARS-CoV-2 antibody (IgG) in different donor types along with vaccinated percent.

### Characteristics of study participants

From February-September 2021, we enrolled 748 CMA service providers (362 HCWs, 205 garments workers, 179 indoor/outdoor patients). Among them, 27.48% were garment workers, 150 (20.11%) hospital staff, 145 (19.44%) doctors, 148 (19.84%) outdoor patients, 67 (8.98%) nurses, and 31 (4.16%) indoor patients. The majority (n=507; 67.96%) were males. In the total population, 292 (39.14%) did not receive any COVID-19 vaccine, 223 (29.89%) received the first dose of vaccine, and 231 (30.97%) received both doses of the vaccine. The responses regarding contact with confirmed COVID-19 cases were: yes (342; 47.17%), no (307; 42.34%), and unknown (76; 10.48%). One hundred and ninety-seven (32.35%) participants had pre-existing medical conditions **(Table 2)**.

**Table 2:** Baseline characteristics of study participants

| **Table 2:** Baseline characteristics of study participants |  |  |  |  |
| --- | --- | --- | --- | --- |
| **Variables** | **Level** | **Total population** | **Known positives (RT-qPCR positive)** | **Asymptomatic** |
| Donor type | Doctor | 145 (19.44) | 40 (35.40) | 85 (16.13) |
|  | Nurse | 67 (8.98) | 19 (16.81) | 43 (8.16) |
|  | Hospital staff | 150 (20.11) | 27 (23.89) | 109 (20.68) |
|  | Indoor patient | 31 (4.16) | 2 (1.77) | 26 (4.93) |
|  | Outdoor patient | 148 (19.84) | 21 (18.58) | 109 (20.68) |
|  | Garments worker | 205 (27.48) | 4 (3.54) | 155 (29.41) |
| Gender | Male | 507 (67.96) | 73 (65.18) | 362 (68.69) |
|  | Female | 239 (32.04) | 39 (34.82) | 165 (31.31) |
| Age (year) | 19 to 29 | 201 (26.91) | 15 (13.27) | 149 (28.27) |
|  | 30 to 35 | 184 (24.63) | 30 (26.55) | 123 (23.34) |
|  | 36 to 44 | 180 (24.10) | 34 (30.09) | 123 (23.34) |
|  | 45 to 84 | 182 (24.36) | 34 (30.09) | 132 (25.05) |
| Vaccination | No | 292 (39.14) | 11 (9.82) | 222 (42.13) |
|  | Only 1<sup>st</sup> dose | 223 (29.89) | 38 (33.93) | 153 (29.03) |
|  | Both doses | 231 (30.97) | 63 (56.25) | 152 (28.84) |
| Days passed after 1<sup>st</sup> dose of vaccine | 14 to 30 days | 45 (24.06) | 8 (25.81) | 30 (23.08; 16.1-31.3) |
|  | 31 to 60 days | 142 (75.94) | 23 (74.19) | 100 (76.92) |
| Days passed after 2<sup>nd</sup> dose vaccine | 14 to 60 days | 19 (8.26) | 6 (9.38) | 12 (8.00) |
|  | 61 to 120 days | 86 (37.39) | 20 (31.25) | 60 (40.00) |
|  | 120 to 180 days | 125 (54.35) | 37 (59.38) | 78 (52.00) |
| Days between PCR test and antibody test | 21 to 60 days | - | 17 (15.60) | - |
|  | 61 to 120 days | - | 16 (14.68) | - |
|  | 121 to 180 days months | - | 23 (21.10) | - |
|  | >180 days | - | 53 (48.62) | - |
| Contact with confirmed case | Yes | 342 (47.17) | 79 (71.17) | 230 (45.19) |
|  | No | 307 (42.34) | 17 (15.32) | 232 (45.58) |
|  | Don't know | 76 (10.48) | 15 (13.51) | 47 (9.23) |
| Family member | 1 to 3 | 186 (26.23) | 31 (29.52) | 130 (25.79) |
|  | 4 to 6 | 443 (62.48) | 64 (60.95) | 321 (63.69) |
| | $\geq 7$ | 80 (11.28) | 10 (9.52) | 53 (10.52) |
| Taking immunosuppressive drugs | Yes | 15 (2.13) | 7 (6.42) | 8 (1.63) |
|  | No | 688 (97.87) | 102 (93.58) | 484 (98.37) |
| Comorbidities | Yes | 197 (32.35) | 38 (37.25) | 291 (68.79) |
|  | No | 412 (67.65) | 64 (62.75) | 132 (31.29) |

### SARS-CoV-2 antibody titer

Indoor/outdoor patients had the highest mean titer of 197.18 DU/mL, followed by HCWs (163.30 DU/mL) and garment workers (77.05 DU/mL) (*p* <0.001). The level (mean) of IgG-spike antibodies in both dosage vaccine recipients was higher (255.46 DU/mL) than in those who received one (159.08 DU/mL) or no doses (53.71 DU/mL) of the vaccine (*p* <0.001). When the participants had a contact with confirmed cases had a mean titer of 170.89 DU/mL, not known had a titer of 160.05 DU/mL, and in case of noncontact 116.45 DU/mL (*p* <0.001). The mean titer of different age groups was statistically significant; nevertheless, we removed this variable from further analysis to minimize the bias due to vaccination strategy followed in Bangladesh (priority given to aged); details in **Table 3**. The changes in mean titer of IgG antibody across different time intervals of intervention (one and both doses of vaccination) is illustrated in **Figure 2**.

**Table 3:**
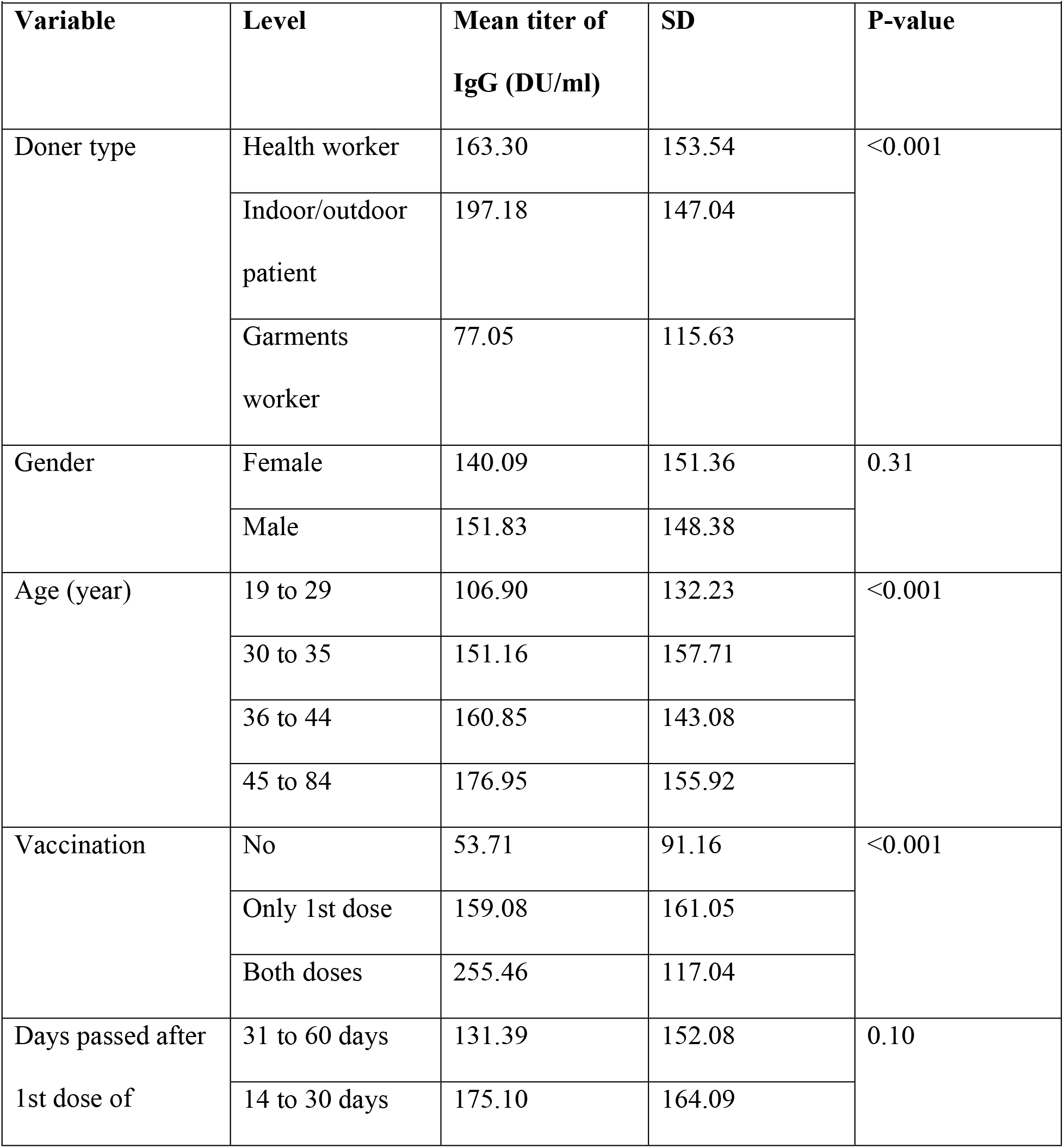
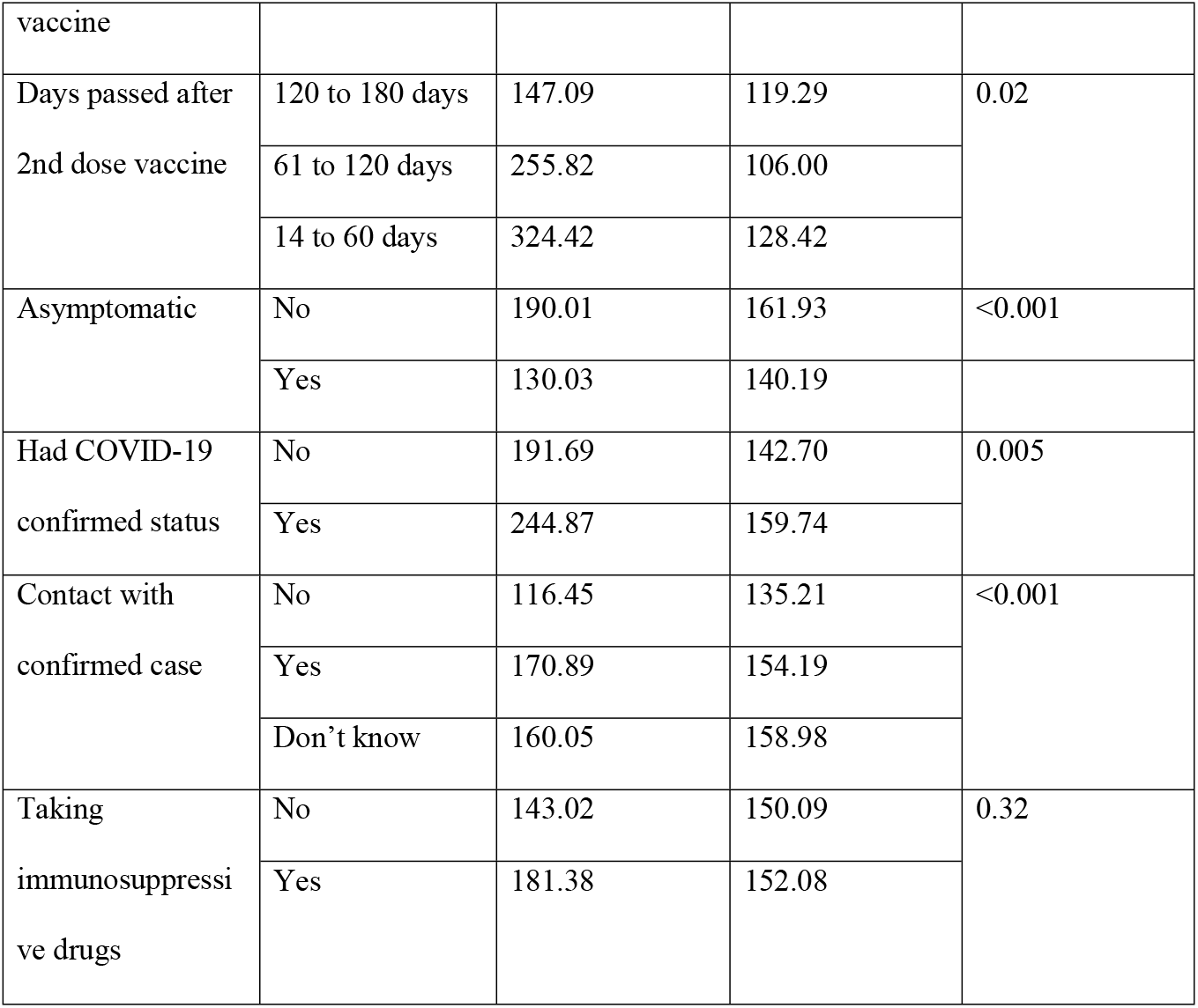
Univariable analysis (ttest, one way ANOVA) to evaluate the mean difference of quantity of anti-SARS-CoV-2 antibody in serum samples

**Figure 2:**
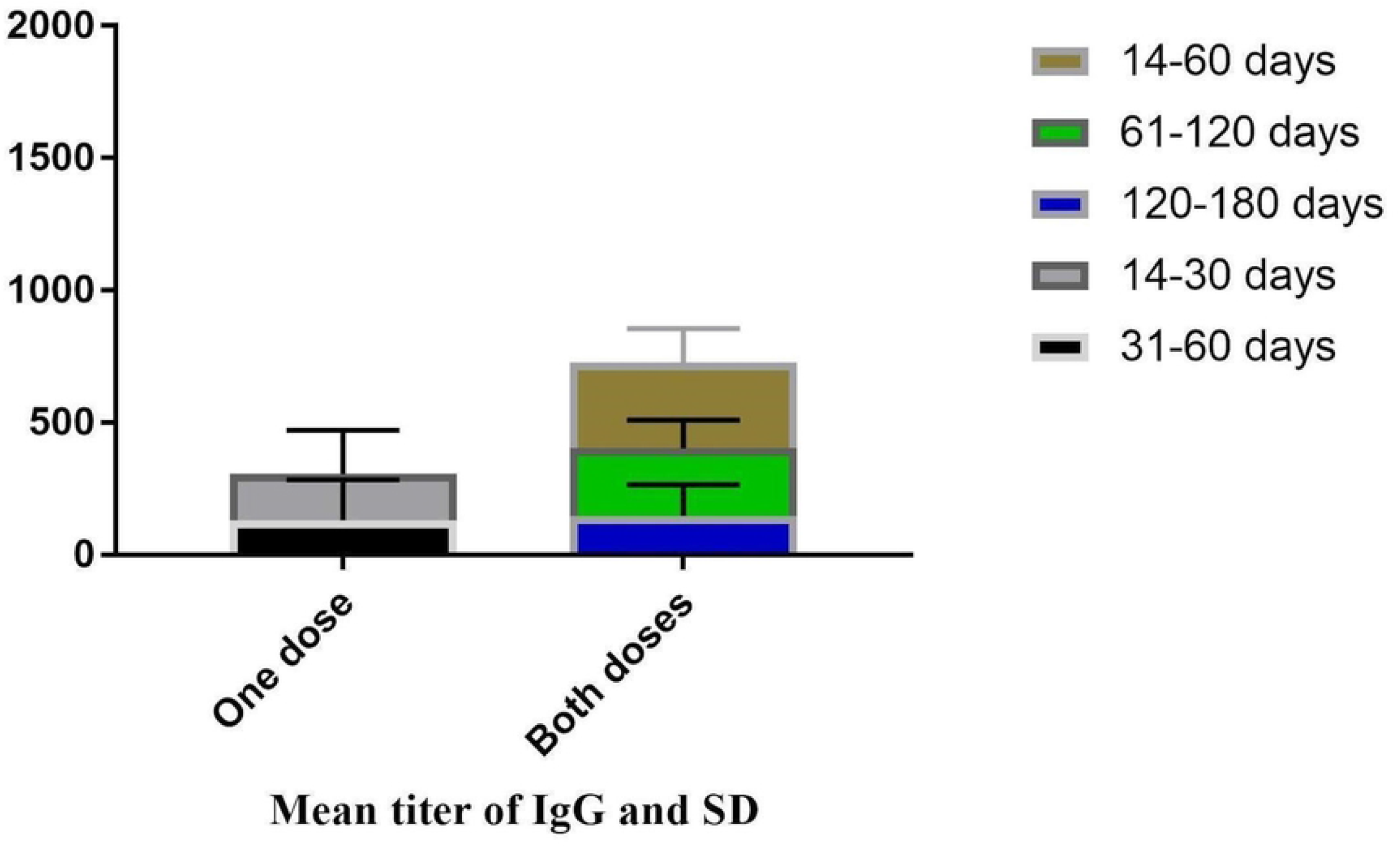
Evaluation of time effects of vaccines mean difference of quantitative anti-SARS-CoV-2 antibody (IgG) in serum samples.

### Risk factor analysis

#### Univariable analysis (χ^2^ test, logistic regression) to evaluate the association of different variables with the seroprevalence of anti-SARS-CoV-2 antibody

Indoor/outdoor patients amongst the different donor groups had a positivity rate of 81.37% (144 of 179) compared to 68.99% (248 of 362) in the HCWs and 50.56% in the garments workers (104 of 205); the difference was statistically significant (*p*< 0.001). Both doses of vaccine receivers showed significantly (*p*<0.001) higher seropositivity than one dose or no vaccine receivers. Similarly, contact with confirmed COVID-19 cases showed a higher odd of being seropositive as compared to noncontact (*p*=0.01) [OR= 1.59] **(Table 4)**.

**Table 4:** Univariable analysis (χ^2^ test, logistic regression) to evaluate the association of different variables with seroprevalence of anti-SARS-CoV-2 antibody

| **Table 4:** Univariable analysis ($\chi^2$ test, logistic regression) to evaluate the association of different variables with seroprevalence of anti-SARS-CoV-2 antibody | | | | | |
| --- | --- | --- | --- | --- | --- |
| **Variable** | **Level (n)** | **Presence of IgG** | **TP (95% CI of TP) \*\*** | **OR** | **P-value** |
| Donor type | Health worker (362) | 248 | 68.99 (63.8-73.7) | Ref. | <0.001 |
|  | Indoor/outdoor patient (179) | 144 | 81.37 (74.7-86.7) | 1.8 |  |
|  | Garments worker (205) | 104 | 50.56 (43.5-57.5) | 0.47 |  |
| Gender | Female (239) | 151 | 63.47 (56.9-69.5) | Ref. | 0.15 |
|  | Male (507) | 347 | 68.92 (64.6-72.9) | 1.26 |  |
| Age (year) | 19 to 29 (201) | 114 | 56.76 (49.5-63.6) | Ref. | 0.002 |
|  | 30 to 35 (184) | 119 | 65.01<br>(57.6-71.8) | 1.39 |  |
|  | 36 to 44 (180) | 132 | 73.99<br>(66.8-80.1) | 2.09 |  |
|  | 45 to 84 (182) | 133 | 73.73<br>(66.6-79.8) | 2.07 |  |
| Vaccination | No (292) | 131 | 44.47<br>(38.6-50.4) | Ref. | <0.001 |
|  | Only 1st dose<br>(223) | 137 | 61.66<br>(54.8-68.0) | 1.95 |  |
|  | Both doses<br>(231) | 229 | 100<br>(98.4-100.0) | 140.72 |  |
| Days passed after<br>1st dose of<br>vaccine | 31 to 60 days<br>(142) | 79 | 55.64<br>(47.1-63.8) | Ref. | 0.29 |
|  | 14 to 30 days<br>(45) | 29 | 64.78<br>(49.6-77.5) | 1.44 |  |
| Days passed after<br>2nd dose vaccine | 120 to 180<br>days (125) | 123 | 99.9 (95.7-100) | - | - |
|  | 61 to 120 days<br>(86) | 86 | 100<br>(97.2-100) | - |  |
|  | 14 to 60 days<br>(19) | 19 | 100<br>(84.2-100) | - |  |
| Asymptomatic | No (220) | 160 | 73.36<br>(66.9-79.03) | Ref. | 0.13 |
|  | Yes (528) | 355 | 67.66<br>(63.4-71.68) | 0.76 |  |
| Had COVID-19 confirmed status | No (144) | 119 | 83.65<br>(76.3-89.1) | Ref. | 0.66 |
|  | Yes (113) | 91 | 81.46<br>(72.9-87.9) | 0.86 |  |
| Contact with confirmed case | No (307) | 187 | 61.11<br>(55.3-66.6) | Ref. | 0.01 |
|  | Yes (342) | 244 | 71.93<br>(66.7-76.6) | 1.59 |  |
|  | Don't know (76) | 49 | 64.81<br>(53.1-75.0) | 1.16 |  |
| Taking immunosuppressive drugs | No (688) | 447 | 65.32<br>(61.5-68.9) | Ref. | 0.20 |
|  | Yes (15) | 12 | 80.91<br>(54.7-94.3) | 2.15 |  |

\*\*TP=True prevalence

#### Multivariable analysis (logistic regression) to determine the potential factors associated with SARS-CoV-2 antibody-positive status in the study area

The multivariable logistic regression model identified two potential factors that might be influencing the seropositivity of SARS-CoV-2 antibodies in the studied population. The chance of being seropositive was 2.22 times higher in indoor/outdoor patients (*p*= 0.002) and 1.69 times for garments workers than HCWs (*p*= 0.01). Further, both doses of vaccine receivers had a higher chance of being positive (OR=174.02) than one dose (OR=2.34) or none dose receivers, and the difference was statistically significant (*p*< 0.001) (**Table 5)**.

**Table 5:** Output from the final multivariable logistic regression model showing the adjusted effect of potential factors on the seroprevalence of anti-SARS-CoV-2 antibody

|  |  |  |  |  |
| --- | --- | --- | --- | --- |
| **Variable** | **Level** | **OR** | **95% CI** | **P-value** |
| Doner type | Health worker | Ref. |  |  |
|  | Indoor/outdoor patient | 2.22 | 1.33-3.68 | 0.002 |
|  | Garments worker | 1.69 | 1.09-2.62 | 0.01 |
| Vaccination | No | Ref. |  |  |
|  | Only 1st dose | 2.34 | 1.56-3.50 | <0.001 |
|  | Both doses | 174.02 | 41.46-730.40 | <0.001 |

## Discussion

The overall adjusted seroprevalence estimate of SARS-CoV-2 antibodies was 66.99% (95% CI: 63.40%-70.4%) in CMA in this research which is slightly higher than a previous finding (64.1%) using an immunoassay test to detect antibodies in the Sitakunda sub-district (Chattogram district) of Bangladesh from March to June 2021 [**36**]. Another research conducted by icddr’b between October 2020 and February 2021 found a lower (55%) estimate in Chattogram than ours. During the same study period, however, the adjusted seroprevalence in Dhaka, the capital of Bangladesh, was 71% [**37**]. Thus, based on several investigations, it can be assumed that seropositivity in Chattogram has been progressively increasing over time. The prevalence might have increased due to either high infection levels or a positive response to the national immunization campaign in its early phases [**38**]. According to the findings, 68.99% HCWs and 81.37% indoor/outdoor patients were seropositive. Indoor and outdoor patients were more likely than health professionals to be seropositive, possibly due to the combined effect of lack of awareness and knowledge about COVID-19 among some of them and effect of vaccination as they might be were composed of a mixed population of lower to upper socio-economic status and with different educational levels. Tripathi et al., 2020 reported that HCWs were more educated of COVID-19 symptoms, incubation time, problems in high-risk patients, and had greater access to therapy than other residents (non HCWs) [**39**]. In Navi Mumbai in May 2021, serosurveillance of anti-SARS-Cov-2 antibodies among essential workers revealed that police personnel had a 72% seropositivity rate, whereas HCWs had a 48% positivity rate [**40**]. Moreover, we observed that, among the garment workers, just under 20% received vaccines and just above 50% were seropositive, which might have majorly been achieved from natural infections (**Figure 1**). It might indicate their lack of awareness about disease transmission and vaccination.

We found that the IgG antibody was produced in 61.66% of the participants who received the first dose of COVID-19 vaccination. This number increased to 100% among individuals who received a second dose. In a study by Bayram et al., 2021, HCWs’ seropositivity rates after the first and second doses of CoronaVac vaccination were found to be 77.8% and 99.6%, respectively [**41**]. Subsequently, when we quantified the antibody titer, we observed it higher in those who received the second dose than in those who received just the first. Detection of highly avid anti-S1/-RBD IgG, independent of the causal mechanism, is seen as a very positive indication and indicator of enhanced humoral immunity [**42**].

Human coronavirus infection may not always result in long-lasting antibody responses, with antibody titers dropping over time [**43**]. The waning of antibody responses is an essential element to consider while developing a coronavirus vaccine [**44**]. Our study showed that by the second month following the initial dose, the mean IgG titer in the body had dropped by nearly 25%. However, the antibody’s propensity to deteriorate with time was noteworthy. This study revealed that the available mean antibody titers that remained after two months of receiving the second dose had dropped by roughly 21% by the fourth month, and within the sixth month the mean antibody titer was 147.09 DU/mL. So, it can be assumed that the body still retained considerable antibodies against COVID-19 six months after receiving the second dose vaccine, though the threshold level to prevent the virus is not known.

The underreporting of SARS-CoV-2 infection cases makes it difficult to assess the actual infection burden. Limited testing, flaws in the reporting infrastructure, and a substantial proportion of asymptomatic infections contribute to the underreporting [**45**]. Asymptomatic carriers spread COVID-19, but the clinical characteristics, viral dynamics, and antibody responses of these individuals are unknown [**46**]. According to our findings, 67.66% of the asymptomatic population was seropositive where only 29.03 % of asymptomatic individuals received the first dose of COVID-19 vaccine, and 28.84 % received the second dose too. According to various population-based studies, a considerable majority of seropositive people were asymptomatic or had no known encounter with a COVID-19 patient [**47–49**]. Meanwhile, the observation that asymptomatic people had lower mean IgG levels than symptomatic people back up previous findings that asymptomatic carriers have a lesser humoral immune response to COVID-19 infection [**47**,**50**]. The study also revealed that people aged above 35 had a greater seroprevalence. Higher seroprevalence among adults could be associated with increased vaccination exposure. On January 26 of this year, the government began accepting registrations for the COVID-19 vaccine for persons aged 55 and up in the country [**51**]. In the second phase, the age limit was dropped to 40 years or more, and vaccination of youngsters aged 12-17 has recently begun in the country [**52**].

The latest and more deadly SARS-CoV-2 viral strains, as well as the possibility of losing immunity with time after vaccination, have prompted health professionals to consider the need for boosters. Research on threshold titers giving protection and time intervals of declining immunity post-immunization for low-middle-income nations like Bangladesh are essential before launching further booster doses. An important application of serological tests is to determine the antibody responses generated upon SARS-CoV-2 infection and vaccination [**53**]. The continuation of this study on those who received the second dose more than six months ago will provide an appropriate booster interval, risk population category, and overview of herd immunity. According to a recent study conducted in the greater Chattogram division it is evident that administering the first dose (Oxford-AstraZeneca) vaccine significantly reduces health risk during the COVID-19 infection phase [**54**]. So, it is evident that similar research is clamoring for justifications for booster administration. Additionally, more research is required to assess the efficacy of booster doses. Government and healthcare professionals must adopt COVID-19 vaccine booster dose utilization guidelines that consider the risks of fading immunity, new virus strains, and prioritizing vulnerable groups.

Our study has several limitations, such as the fact that we only collected samples from hospitals and the garment industry, but the results would be more representative of the community if we included other groups. We could not compare immunological responses produced by different COVID-19 vaccine brands at the same post-vaccination interval since distinct COVID-19 vaccines were licensed and supplied to CMA at different times. We did not reveal the type and name of COVID-19 vaccines, whereas a sufficient fraction was not covered under the vaccination program, and we were concerned about an infodemic.

## Data Availability

All relevant data are within the manuscript and its Supporting Information files

## Acknowledgment

The authors acknowledge Bangladesh Institute of Tropical and Infectious Diseases (BITID), Chattogram General Hospital, Chittagong Medical University, Medical Center (Pvt.), Imperial Hospital Limited, Chattogram Asian Apparels Limited, Clifton Garments Limited, Bangladesh Garment Manufacturers and Exporters Association (BGMEA) Chattogram, and BGMEA Hospital and Diagnostic Center for providing samples and administrative support. The laboratory facilities were supported by the Dept. of Pathology and Parasitology, CVASU. The authors sincerely acknowledge the One Health Institute, CVASU for logistic support.

## Authors Contribution

**JA:** Conceptualization, methodology, project proposal writing, administration, data curation, writing-original draft, review and editing. **MSI:** Conceptualization, methodology, project administration, laboratory tasks, data visualization, writing-original draft, review and editing. **MTUQ:** Sample collection, data curation. **AD:** Sample collection, data curation. **FMYH:** Laboratory tasks. **MSI:** Sample collection, data curation. **TR:** Sample collection, data curation. **SD:** Data curation, writing review and editing. **MAHC:** Conceptualization, investigation, methodology, supervision, and reviewing-final manuscript. **GBD:** Project investigation, fund acquisition, and reviewing-final manuscript. **SC:** Conceptualization, investigation, methodology, supervision, data-validation, analysis, visualization, writing-original draft, and reviewing-final manuscript.

